# The individual and joint associations of alcohol use and cigarette smoking in adolescence and early adulthood with psychological distress in midlife – a multicohort study

**DOI:** 10.1101/2025.07.11.25330860

**Authors:** Noora Berg, Jenna Grundström, Sari Aaltonen, Antti Latvala, Jouko Miettunen, Olli Kiviruusu

**Affiliations:** Department of Healthcare and Social Welfare, Finnish Institute for Health and Welfare, Helsinki, Finland; Institute for Molecular Medicine Finland, FIMM, HiLIFE, University of Helsinki, Helsinki, Finland; Institute of Criminology and Legal Policy, University of Helsinki, Helsinki, Finland; Research Unit of Population Health, University of Oulu, Oulu, Finland; Medical Research Center Oulu, Oulu University Hospital and University of Oulu, Oulu, Finland

**Keywords:** alcohol, life course, mental health, smoking, tobacco

## Abstract

**Background:** Alcohol and tobacco use are associated with later mental health and psychological well-being, yet the associations of combined use of alcohol and tobacco with psychological well-being throughout the life span remain understudied. The aim of this study is to examine the separate and combined associations of heavy episodic drinking (HED) and daily cigarette smoking in adolescence and early adulthood with midlife psychological distress across four prospective cohort studies from Finland and Sweden.

**Methods:** Data for this prospective multicohort study were drawn from four longitudinal studies: the ‘TAM’ cohort (N=1334), the ‘FinnTwin16 Study’ (N=4409), and the ‘Northern Finland Birth Cohort 1966’ (NFBC1966, N=7147) from Finland, and the ‘Individual Development and Adaptation Study’ (IDA, N=514) from Sweden. HED was measured as drunkenness or using 60g or more pure alcohol on one occasion, smoking as daily cigarette use, and psychological distress using the 12-item General Health Questionnaire.

**Results:** No clear evidence for individual or combined associations of HED or smoking in adolescence or early adulthood with midlife distress were found. The results were overall systematic across all cohorts.

**Conclusions:** In contrast to prior research based on diagnostic criteria, this multicohort study found no evidence that heavy episodic drinking or daily cigarette smoking in adolescence or early adulthood— individually or combined—are associated with midlife psychological distress when assessed using subclinical symptom measures. These findings, overall consistent across four longitudinal cohorts, suggest that the relationship between early substance use and later mental health may be weaker than previously assumed when studied using non-diagnostic indicators.

## Introduction

Substance use and mental health problems are major public health concerns worldwide (Peacock et al., 2018; Rehm and Shield, 2019). Alcohol and tobacco use are correlated with mental health issues, although there is more evidence for an association between alcohol use and mental health disorders than for subclinical indicators (Puddephatt et al., 2022) and longitudinal findings of smoking and mental health are inconsistent (Fluharty et al., 2017). The causal relationship between substance use and mental health is likely bidirectional: while individuals may use substances like alcohol and tobacco to cope with mental health problems, substance use can, in turn, exacerbate or trigger psychological symptoms (e.g., Bell & Britton, 2014; Fergusson et al., 2009). Alcohol consumption, especially heavy consumption, and daily tobacco use, has been found to be associated with increased risk of mental health problems, such as psychological disorders or depression (de Boer et al., 2023), but some studies have not found this association (Berg et al., 2019; Schulte & Li, 2022). These inconsistencies may be due to differences in how studies account for genetic and environmental confounding factors, or they may result from differing methodologies and measurements used (Ranjit et al., 2019a). Genetically informative studies have concluded that the longitudinal association between substance use and mental health may, for the most part, be explained by environmental and genetic factors (Kuo et al., 2006; Polimanti et al., 2019; Ranjit et al., 2019b).

The use of alcohol and tobacco typically begins in adolescence and peaks in early adulthood (Chassin et al., 2018; Schulenberg et al., 2018), periods characterized by significant developmental changes and vulnerability. Adolescence has been described as a sensitive period characterized by ongoing physical, emotional, and cognitive development, making individuals more susceptible to engaging in risky behaviors such as substance use (Green and Popham, 2017; Jordan and Andersen, 2017). Early adulthood is a time when individuals start gaining more independence and autonomy, which can increase substance use (Schulte and Li, 2022). In most countries, majority is reached at age 18-21 years, which often also marks the legal age to buy alcohol and tobacco. Both adolescence and early adulthood represent critical windows of increased risk for the development of substance use disorders (Jordan and Andersen, 2017). However, it remains poorly known when during the life span substance use shifts from experimenting to a habit (Schulte and Li, 2022). Early initiation of alcohol and tobacco use can disrupt normal developmental processes and establish patterns of behavior that persist into later life. Early substance use can have long-lasting health implications, including continued substance use and dependence (McCambridge et al., 2011). These early behaviors may also have enduring effects on mental health, with alcohol and tobacco use in adolescence or early adulthood acting as risk factors for mental health problems in midlife (e.g., Lee et al., 2022; Trim et al., 2007). Possible mechanisms explaining this longitudinal association have been related to the neurobiological effects of repeated intoxication and withdrawal, as well as effects related to social and economic circumstances (Boden and Fergusson, 2011).

Alcohol and tobacco use are associated with later mental health and psychological well-being (Lee et al., 2022; Skogen et al., 2016), yet the associations of combined use of alcohol and tobacco with psychological well-being throughout the life span remain understudied. These combined associations of alcohol and tobacco use may be linked through social, psychological, and physiological mechanisms. Previous studies have suggested that alcohol and tobacco use act as complementary behaviors, where frequent alcohol use increases the likelihood of frequent smoking – and vice versa (Witvorapong and Vichitkunakorn, 2021). When combined, these behaviors can amplify their longitudinal effects, potentially leading to more severe and persistent mental health challenges in mid-adulthood. Previous studies have mainly examined the accumulated effect of various health behaviors, and found that the more healthy behaviors a person engages in, the higher their life satisfaction and lower psychological distress (Stenlund et al., 2021; Velten et al., 2014). For instance, studies have used composite scores to aggregate multiple health behaviors into a single measure, providing a more holistic view of their combined association with an individual’s well-being. Stenlund et al. (2021), for example, examined the composite score of dietary habits, smoking, alcohol use, and physical activity on life satisfaction in a 9-year follow-up and found that healthier behaviors had a positive association with life satisfaction. However, studies specifically investigating the combined association of alcohol and tobacco use with mental health are still rare. Given the frequent co-use of these substances, further research on their individual and additive associations is needed to understand how they may jointly contribute to long-term psychological outcomes.

Typically, men use alcohol and tobacco in larger amounts and more often than women (Zahn-Waxler et al., 2008), although the gender gap in alcohol and tobacco use is narrowing in younger cohorts (CAN, 2017; Slade et al., 2016). Further, women usually report more mental health symptoms (Zahn-Waxler et al., 2008), suggesting that these gender differences should be taken into account and further studied in substance use and mental health research.

## Aim

This study aims to address the limitations of previous studies by focusing simultaneously on alcohol use and cigarette smoking and exploring substance use in two different life stages in men and women. The focus of this study is on the association between excessive substance use and subsequent mental health problems. The aim is to examine both separate and combined associations of heavy episodic drinking (HED) and daily cigarette smoking in adolescence and early adulthood with midlife psychological distress in four prospective cohort studies.

## Research questions

1. Are heavy episodic drinking and daily cigarette smoking in adolescence and early adulthood associated with midlife psychological distress?

Hypothesis 1: We expect a positive association, though potentially weaker than in studies using diagnostic outcomes, due to our use of subclinical measures.

2. Is the timing and persistence of risky substance use (i.e., adolescence only, early adulthood only, or continued from adolescence to early adulthood) associated with midlife psychological distress?

Hypothesis 2: We expect the strongest associations when substance use begins in adolescence and continues into early adulthood. The associations are stronger in adolescence compared to early adulthood, because substance use is more common/normative in early adulthood.

3. Is there an interaction effect between heavy episodic drinking and daily smoking on midlife psychological distress?

Hypothesis 3: We expect a greater risk for psychological distress when both behaviors co-occur, beyond the sum of their individual effects.

We examine the persistence of these associations across four different cohorts from Finland and Sweden, covering 19 to 36 years of follow-up data. We conduct a two-step individual participant meta-analysis. First, we conduct aligned analyses in each cohort, and then meta-analyse the estimates.

4. Do these associations vary across cohorts from Finland and Sweden?

Hypothesis 4: We expect stronger associations in older cohorts because substance use is an indicator of more deviant behavior in these cohorts (especially in women) as liberalization of alcohol control policies and attitudes began in the 1960s (Härkönen, 2013).

## Materials and methods

### Populations

The present study uses four different follow-up studies: The ‘TAM’ cohort (Berg et al., 2021), the ‘FinnTwin16 Study’ (FT16) (Kaidesoja et al., 2019), and the ‘Northern Finland Birth Cohort 1966’ (NFBC1966) (Nordström et al., 2021; University of Oulu, 1966) from Finland, and the ‘Individual Development and Adaptation Study’ (IDA) (Bergman et al., 2018) from Sweden. Each cohort study has followed participants from adolescence to adulthood/midlife (Figure 1).

**Figure 1.**
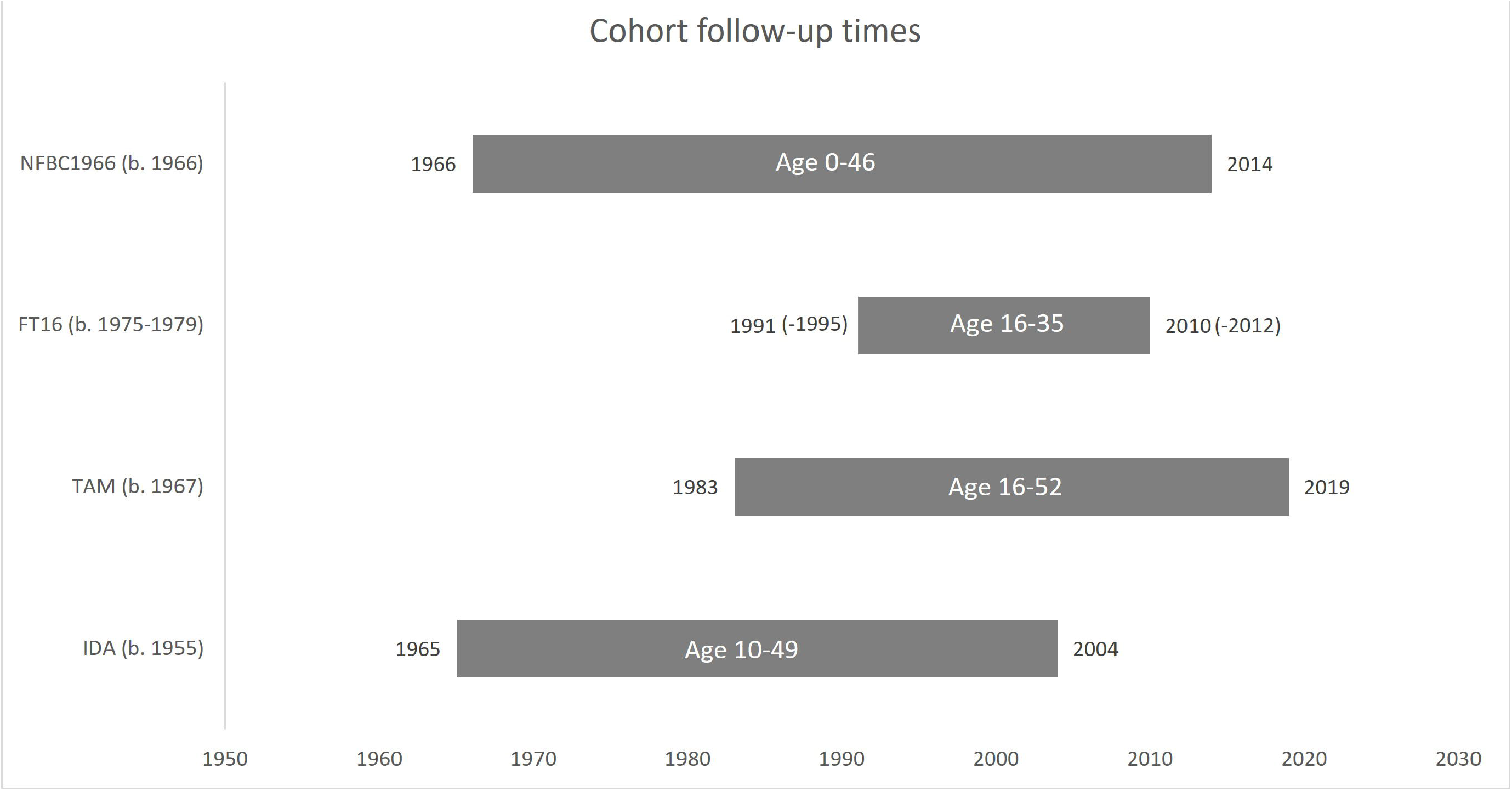
Follow-up times, participants’ birth years, and participants’ ages of the Northern Finland Birth Cohort 1966 (NFBC1966), FinnTwin16 (FT16), TAM cohort, and Individual Development and Adaptation (IDA).

The TAM cohort included all 9^th^ grade (age 16) Finnish-speaking pupils attending comprehensive school in 1983 in Tampere, a city in Southern Finland. In 1983, 2194 pupils (96.7%) completed the questionnaire during the school day, and the participants were then followed up at ages 22, 32, 42, and 52.

FinnTwin16 cohort (FT16) is a follow-up study of Finnish twins born in 1974 ⍰ 1979 (N=5563). The first questionnaires were sent to 16-year-old twins between 1991 and 1995. The follow-up studies were conducted at ages 17, 18, 25, and 35.

The Northern Finland Birth Cohort 1966 study (NFBC1966) includes participants born in 1966 (N=12 058). The data were first collected at the 24^th^ gestational week, and follow-up questionnaires and clinical examinations were conducted at ages 1, 14, 24 (only men), 31, and 46.

The Individual Development and Adaptation (IDA) project started in 1965 when 3^rd^ grade (age 10) students in Örebro Sweden answered the questionnaire. The follow-up questionnaires were conducted at the ages of 13, 15, 16, 17, 18, 19, 26, 43 (women), 47-48 (47 women, 47-48 men), and 49.

We harmonized the developmental timing of measurements so that data collected in adolescence, early adulthood, and mid-adulthood is used, resulting in utilizing data that in the TAM cohort was collected at ages 16, 22, and 42 years, in the FT16 16, 25, and 35 years, in the NFBC1966 14, 24, and 46 years, and in the IDA 14, 27, and 49 years. Because in the IDA cohort, data on midlife psychological distress and adolescent mental health symptoms were available only for females, men were excluded in this cohort. In the NFBC1966, early adulthood substance use is analyzed only in men, because no data is available for females at age 24. The data is described in more detail in Supplementary Table 1.

### Measures

Original and harmonized variables are described in detail in Supplementary Table 2.

#### Midlife psychological distress

In all four cohorts, midlife psychological distress was assessed using the 12-item General Health Questionnaire (GHQ-12) (Goldberg, 1972). The GHQ-12 includes 12 questions assessing mental health problems over the past few weeks, e.g., the ability to make decisions or feeling unhappy and depressed. Items are scored on a four-point scale (from 0 to 3). A higher total score indicates poorer psychological distress. We used a cut-off of 3 or more to high scores.

#### Adolescent heavy episodic drinking

Adolescent heavy episodic drinking was obtained from the questions of frequency of (or experiences of) drunkenness (Supplementary Table 2). Heavy episodic drinking was categorized into two classes in each follow-up study: no experiences of drunkenness or only a few times, and several experiences of being drunk.

#### Heavy episodic drinking in early adulthood

Heavy episodic drinking in early adulthood was measured with questions of frequency of drunkenness (TAM, FT16, NFBC1966) and frequency of drinking occasions using 60 grams or more pure alcohol on one occasion (IDA). Based on these questions, a harmonized variable of HED was constructed. In the Finnish cohorts, if the participants reported drunkenness at least monthly, they were defined as ‘heavy episodic drinkers’, and if they reported drunkenness less often, they were classified as ‘not heavy episodic drinkers’. In the IDA cohort, HED was measured by the frequency of drinking occasions and by the average intake of alcoholic drinks on each occasion. Those who reported drinking at least a couple of times monthly and drinking on average 5 or more drinks at one time were defined as ‘heavy episodic drinkers’. Participants who reported drinking less frequently this amount (or not drinking at all) were classified as ‘not heavy episodic drinkers’.

#### Adolescent and early adulthood cigarette smoking

Smoking status was measured from the questions about cigarette smoking habits and was classified as “daily smoking” and “other smoking statuses” (including never, occasionally, and quit smoking). In the IDA cohort, smoking in adolescence and early adulthood was assessed retrospectively at age 43.

#### Control variables

Adolescent/early adulthood psychological symptoms was used to control the baseline level of the studied outcome. In the TAM, IDA, and FT16 cohorts, a list of various available psychological symptoms was combined into a sum score, which was further standardized. In NFBC1966, adolescent symptoms were not available. Parental socioeconomic position was based primarily on occupation (upper non-manual, lower non-manual, manual) and, if not available, on education (high, intermediate, low). In TAM and NFBC1966 adolescents provided the information, and in other cohorts, the parents.

### Statistical analyses

Logistic regression analyses were performed to examine research questions 1-3 across four different cohorts. The nature of the twin data, i.e., within-pair/family-level clustering, was accounted for in these analyses by using generalized mixed modelling. First, a question about the individual associations between substance use during adolescence and early adulthood and psychological distress in midlife was examined by substance and age (adolescent alcohol use, early adulthood alcohol use, adolescent smoking, early adulthood smoking). Unadjusted models were analysed first, and then in the adjusted models, parental socioeconomic position and psychological symptoms in adolescence or early adulthood (depending on the analyzed age stage) were mutually adjusted for.

To examine research question 2, i.e., the timing and persistence, we formed a 4-class variable by substance: i) no substance use in adolescence and early adulthood (reference category), ii) only adolescence substance use, iii) only early adulthood substance use, iv) substance use in both adolescence and early adulthood.

To explore research question 3, i.e., the combined association between heavy episodic drinking and smoking on midlife psychological distress, we formed 4-class variables by age phase: i) no HED nor smoking (reference category), ii) only HED, iii) only smoking, and iv) both.

We used the Relative Excess Risk due to Interaction (RERI) method to examine the potential additive interaction in research questions 2 and 3. This method is particularly useful for assessing whether the combined effect of two risk factors (or age phases) is greater than the sum of their individual effects. RERI⍰= 0 indicates the absence of an interaction or mere additivity; RERI ≥ 0 indicates a positive interaction or an effect that is more than additivity (i.e., having the two risk factors constitutes a more pronounced risk for an outcome than what would have been expected by just summing the separate risks of the two factors), and RERI < 0 indicates a negative interaction or less than additivity. The 95% CIs for the RERIs were calculated as suggested by Knol et al. (2011).

First, aligned analyses were conducted in all cohorts. Subsequently, research question 4 was addressed by pooling the cohort-specific estimates using multivariate random-effects meta-analysis. Heterogeneity in the effect sizes was examined using the *I*^*2*^ estimates. The data were analyzed using IBM SPSS Statistics 29.0, and meta-analysis was conducted using Stata 18. Women and men were examined separately. A p-value of p < 0.05 was used as a threshold to determine statistical significance. Adolescence was measured two years earlier (age 14 vs. 16) in the NFBC1966 compared to other cohorts. We conducted sensitivity analyses, repeating the analyses in the TAM cohort using age 15 measures of HED and smoking to gain a more similar measurement age with NFBC1966. This data was available for most participants in the TAM cohort.

## Results

The total sample included 13 404 participants. Descriptive statistics of the distribution of study variables by cohort are shown in Table 1 and Supplementary Table 3. The proportion of those with high psychological distress varied between cohorts, 15.2 ⍰ 29.8% in women and 10.8 ⍰ 21.7% in men. In general, substance use was more common in early adulthood than in adolescence. The proportion of participants with low socioeconomic family background ranged between 19 – 65% in the cohorts.

**Table 1.**
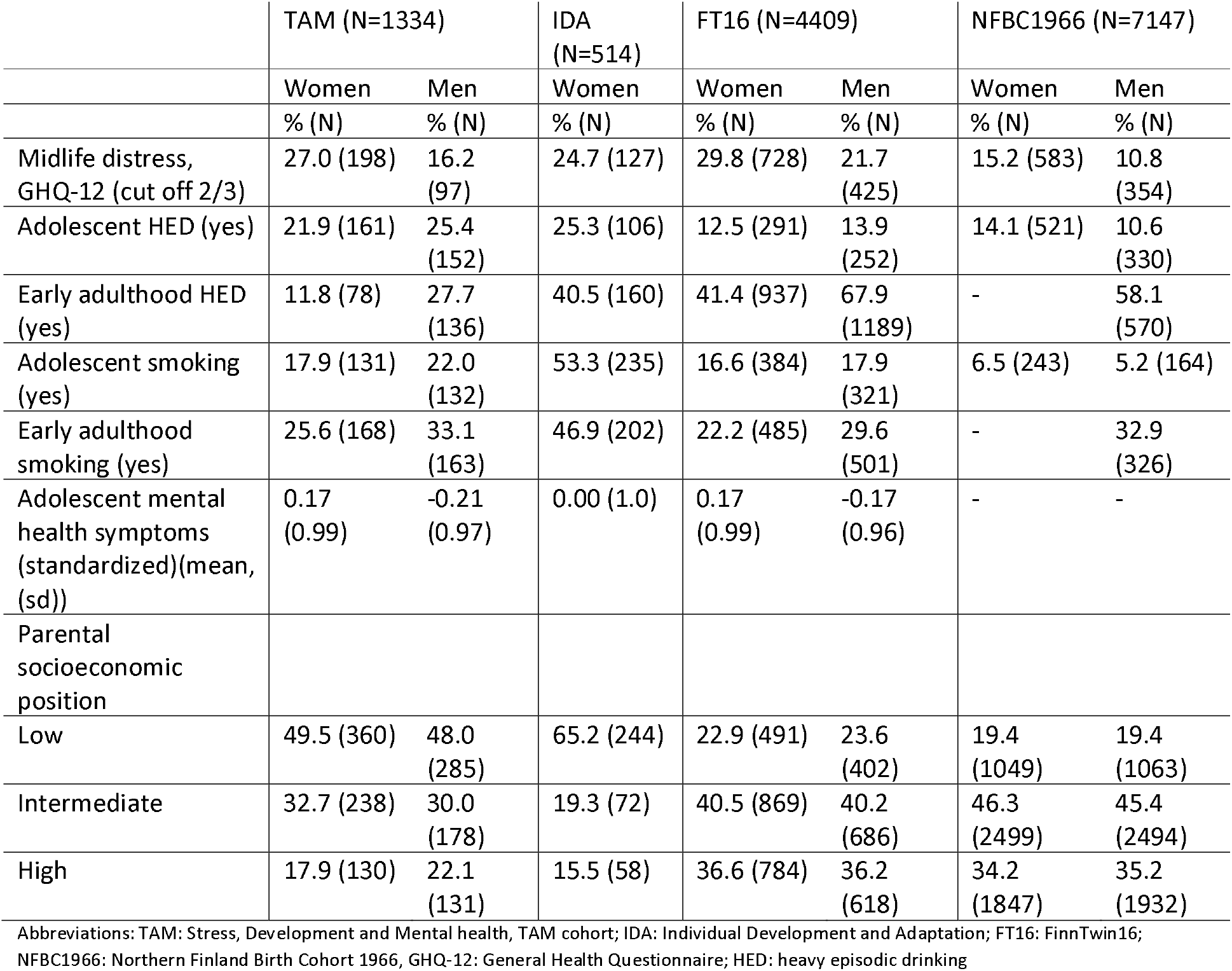
Distributions of study variables in four cohorts.

### Individual associations of HED and smoking with GHQ-12

The univariate logistic regression analyses were used to examine how adolescent and early adulthood heavy episodic drinking and daily cigarette smoking were associated with midlife psychological distress across all four cohorts (Table 2, Figure 2, Figure 3). Among women, the results consistently indicated a lack of clear evidence for associations between adolescent and early adulthood heavy episodic drinking or smoking and midlife psychological distress (Table 2). After adjusting for parental SEP and early adulthood mental health status, a statistically significant association was found in the women of the TAM cohort between early adulthood smoking and midlife distress, and when further studied (results not shown) suggesting that smoking in early adulthood was associated with less psychological distress in midlife in those women who had high symptoms in adolescence but not among those with low symptoms at baseline. Among men, there was no clear evidence of associations between HED or smoking in adolescence or early adulthood and midlife psychological distress across any of the cohorts (Table 2). Meta-analysis (Figure 3) suggested no pooled associations for these relationships in men. *I*^*2*^ estimates showed low to substantial heterogeneity (0 ⍰ 66%), but the Q-values were mainly statistically non-significant (p>0.05), suggesting homogeneity across studies. In women, there was some heterogeneity in the association between early adulthood HED and midlife psychological distress (*I*^*2*^=67.0%, p=0.048).

**Figure 2.**
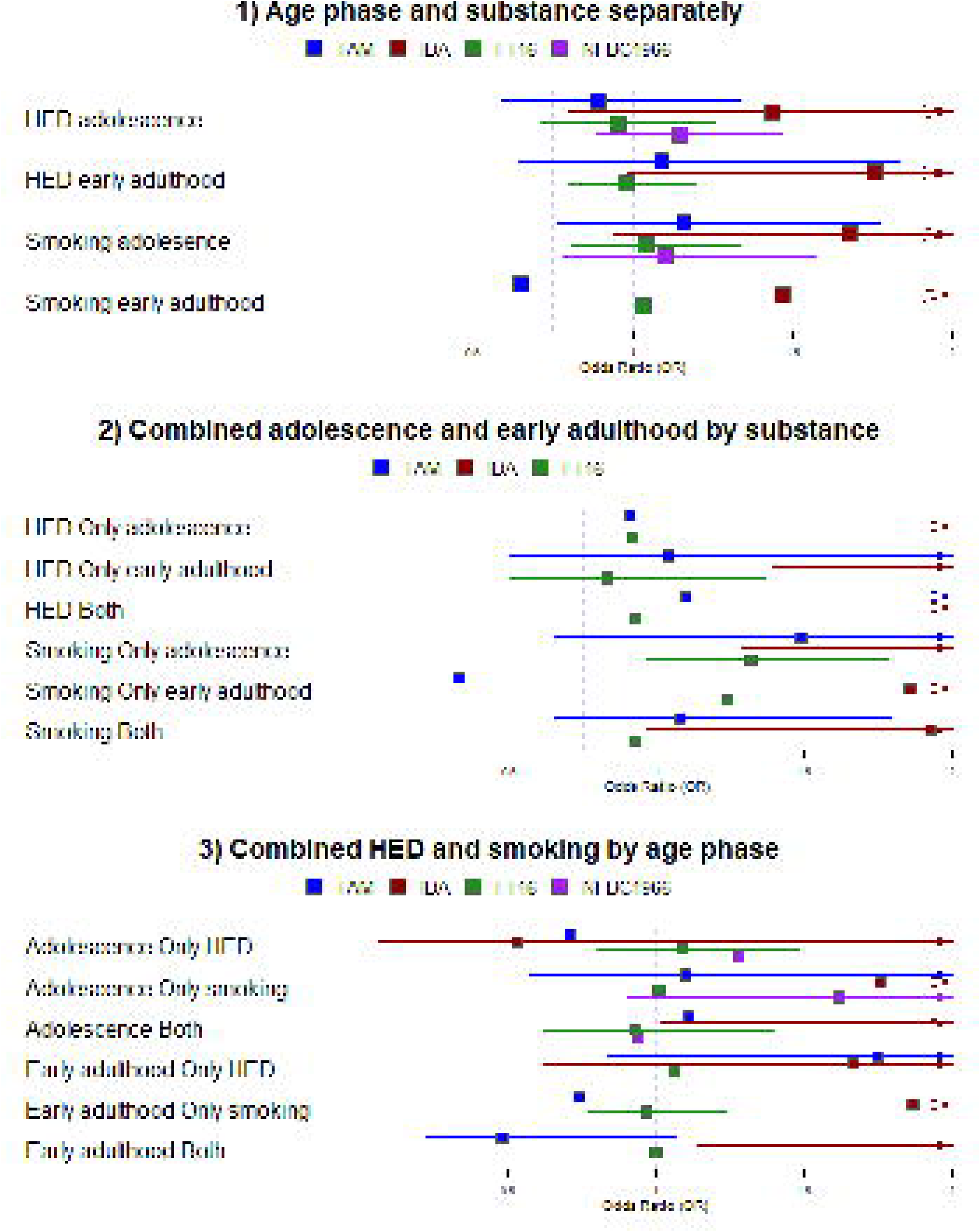
Forest plot of odds ratios (95% confidence interval) for the associations between 1) age phase and substance separately, 2) combined age phase by substance, 3) combined substance by age phase and midlife psychological distress in a random-effect meta-analysis framework in women. TAM: Stress, Development and Mental health, TAM cohort; IDA: Individual Development and Adaptation; FT16: FinnTwin16; NFBC1966: Northern Finland Birth Cohort 1966, GHQ-12: General Health Questionnaire; HED: heavy episodic drinking.

**Table 2.**
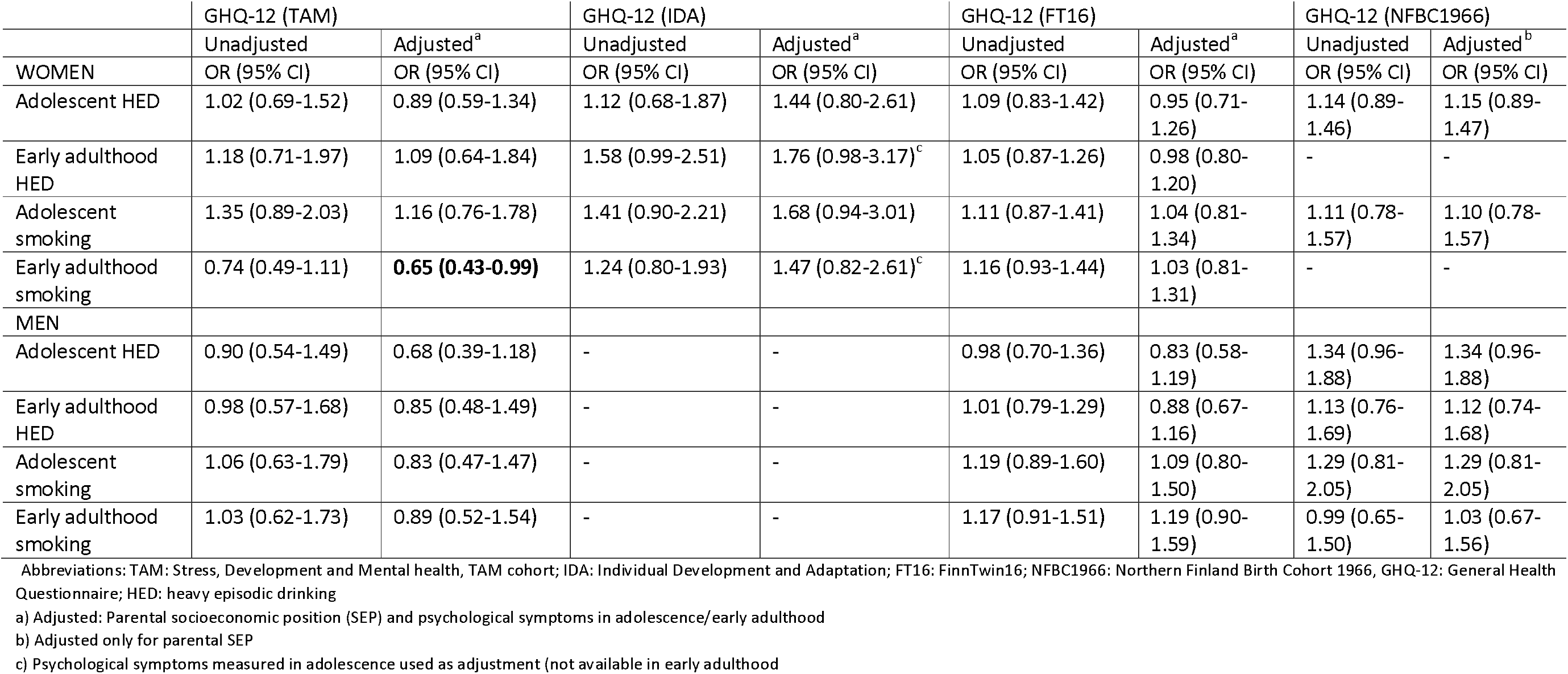
Unadjusted and adjusted logistic regression (and generalized mixed models in FT16) analyses of the associations between adolescent/early adulthood heavy episodic drinking/smoking and midlife psychological distress in women and men.

|  | GHQ-12 (TAM) |  | GHQ-12 (IDA) |  | GHQ-12 (FT16) |  | GHQ-12 (NFBC1966) |  |
| --- | --- | --- | --- | --- | --- | --- | --- | --- |
|  | Unadjusted | Adjusted<sup>a</sup> | Unadjusted | Adjusted<sup>a</sup> | Unadjusted | Adjusted<sup>a</sup> | Unadjusted | Adjusted<sup>b</sup> |
| WOMEN | OR (95% CI) | OR (95% CI) | OR (95% CI) | OR (95% CI) | OR (95% CI) | OR (95% CI) | OR (95% CI) | OR (95% CI) |
| Adolescent HED | 1.02 (0.69-1.52) | 0.89 (0.59-1.34) | 1.12 (0.68-1.87) | 1.44 (0.80-2.61) | 1.09 (0.83-1.42) | 0.95 (0.71-1.26) | 1.14 (0.89-1.46) | 1.15 (0.89-1.47) |
| Early adulthood HED | 1.18 (0.71-1.97) | 1.09 (0.64-1.84) | 1.58 (0.99-2.51) | 1.76 (0.98-3.17)<sup>c</sup> | 1.05 (0.87-1.26) | 0.98 (0.80-1.20) | - | - |
| Adolescent smoking | 1.35 (0.89-2.03) | 1.16 (0.76-1.78) | 1.41 (0.90-2.21) | 1.68 (0.94-3.01) | 1.11 (0.87-1.41) | 1.04 (0.81-1.34) | 1.11 (0.78-1.57) | 1.10 (0.78-1.57) |
| Early adulthood smoking | 0.74 (0.49-1.11) | **0.65 (0.43-0.99)** | 1.24 (0.80-1.93) | 1.47 (0.82-2.61)<sup>c</sup> | 1.16 (0.93-1.44) | 1.03 (0.81-1.31) | - | - |
| MEN |  |  |  |  |  |  |  |  |
| Adolescent HED | 0.90 (0.54-1.49) | 0.68 (0.39-1.18) | - | - | 0.98 (0.70-1.36) | 0.83 (0.58-1.19) | 1.34 (0.96-1.88) | 1.34 (0.96-1.88) |
| Early adulthood HED | 0.98 (0.57-1.68) | 0.85 (0.48-1.49) | - | - | 1.01 (0.79-1.29) | 0.88 (0.67-1.16) | 1.13 (0.76-1.69) | 1.12 (0.74-1.68) |
| Adolescent smoking | 1.06 (0.63-1.79) | 0.83 (0.47-1.47) | - | - | 1.19 (0.89-1.60) | 1.09 (0.80-1.50) | 1.29 (0.81-2.05) | 1.29 (0.81-2.05) |
| Early adulthood smoking | 1.03 (0.62-1.73) | 0.89 (0.52-1.54) | - | - | 1.17 (0.91-1.51) | 1.19 (0.90-1.59) | 0.99 (0.65-1.50) | 1.03 (0.67-1.56) |

Abbreviations: TAM: Stress, Development and Mental health, TAM cohort; IDA: Individual Development and Adaptation; FT16: FinnTwin16; NFBC1966: Northern Finland Birth Cohort 1966, GHQ-12: General Health Questionnaire; HED: heavy episodic drinking
a) Adjusted: Parental socioeconomic position (SEP) and psychological symptoms in adolescence/early adulthood
b) Adjusted only for parental SEP
c) Psychological symptoms measured in adolescence used as adjustment (not available in early adulthood)

**Figure 3.**
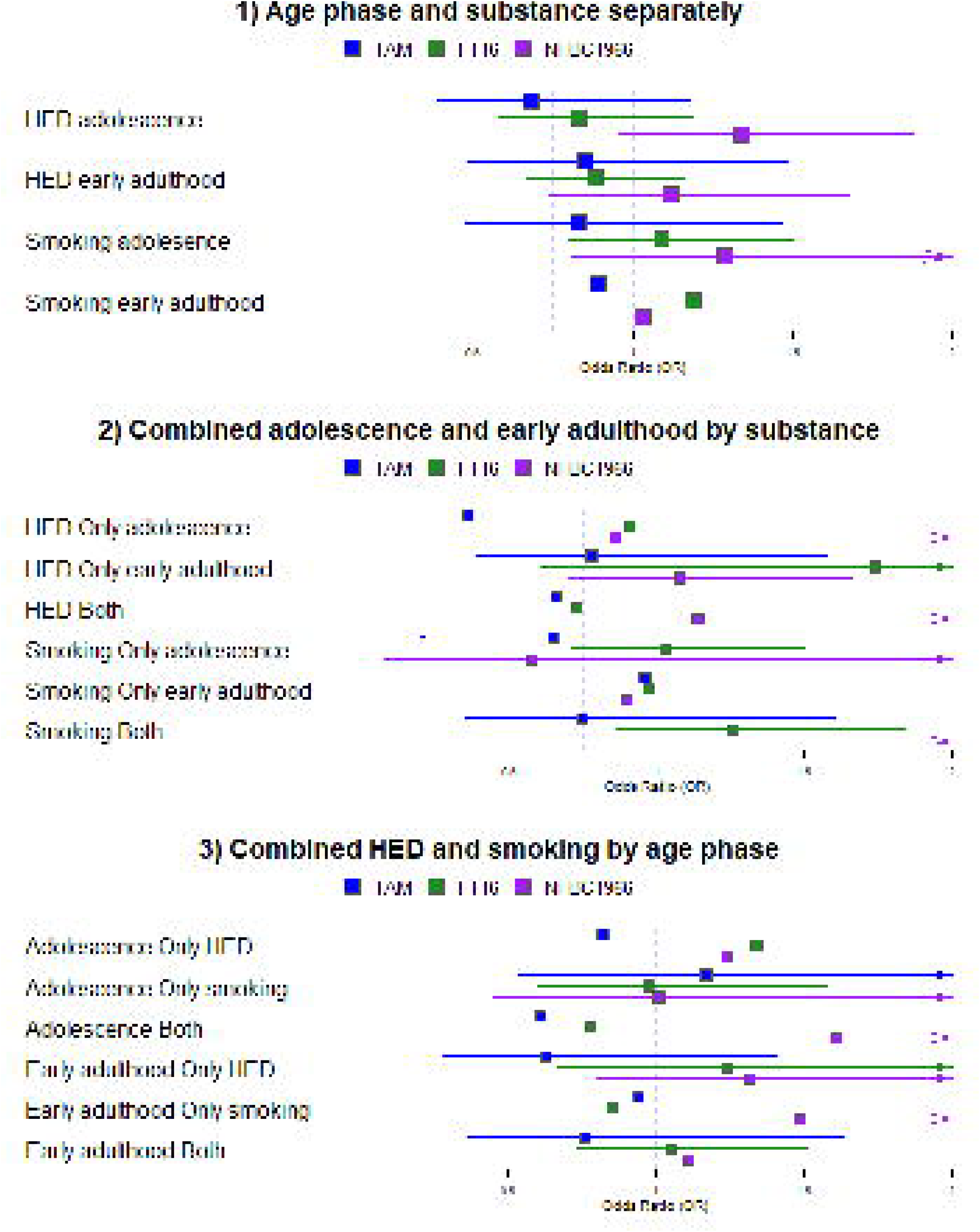
Forest plot of odds ratios (95% confidence interval) for the associations between 1) age phase and substance separately, 2) combined age phase by substance, 3) combined substance by age phase and midlife psychological distress in a random-effect meta-analysis framework in men. TAM: Stress, Development and Mental health, TAM cohort; IDA: Individual Development and Adaptation; FT16: FinnTwin16; NFBC1966: Northern Finland Birth Cohort 1966, GHQ-12: General Health Questionnaire; HED: heavy episodic drinking.

### Differences between age phases

We examined the role of age phase on the associations between HED/smoking and distress (Table 3). There was heterogeneity in the associations for women, with the Swedish cohort indicating a tendency for higher ORs than the Finnish cohorts (HED *I*^*2*^ 73-87%, smoking *I*^*2*^ 69-86%). In the Swedish cohort, associations between adolescence limited HED (OR 2.91, 95% CI 1.08-7.86) and early adulthood limited HED (OR 2.88 (1.39-5.93) were associated with midlife distress. Regarding smoking, association were found for adolescence limited smoking in Swedish women (OR 3.07, 95% CI 1.29-7.35) and women in the TAM cohort (0.,33 (0.16-0.66). In men, the associations were homogenously (HED *I*^*2*^ 0-11%, smoking *I*^*2*^ 0-53%) non-significant, apart from an association between adolescence limited HED and distress in the TAM cohort (OR 0.36, 95% CI 0.13-0.99). Calculating RERIs showed no statistically significant additive interactions, suggesting that engaging in HED/smoking in both adolescence and early adulthood did not constitute a more pronounced association with distress than what would have been expected by just summing the separate associations of the two age phases.

**Table 3.** Logistic regression analyses (and generalized mixed models in FT16) of the association between combined adolescence + early adulthood heavy episodic drinking / smoking and midlife psychological distress.

|  | TAM GHQ-12 | IDA GHQ-12 | FT16 GHQ-12 | NFBC1966 GHQ-12<sup>b</sup> |
| --- | --- | --- | --- | --- |
| **HEAVY EPISODIC DRINKING** |  |  |  |  |
|  | N=656 | N=252 | N=1989 | - |
| **Women** | OR (95%CI) | OR (95%CI) | OR (95%CI) | OR (95%CI) |
| No adol. – no early adult. | 1 | 1 | 1 | - |
| Yes adol. – no early adult. | 0.91 (0.56-1.49) | **2.91 (1.08-7.86)** | 0.92 (0.74-1.14) | - |
| No adol. – yes early adult. | 1.04 (0.50-2.17) | **2.88 (1.39-5.93)** | 0.83 (0.50-1.37) | - |
| Yes adol. – yes early adult. | 1.10 (0.53-2.27) | 2.34 (0.91-6.02) | 0.93 (0.65-1.35) | - |
| RERI<sup>a</sup> (95% CI) | 0.14 (-1.00-1.28) | -2.44 (-6.28-1.40) | 0.19 (-0.36-0.73) | - |
| **Men** | N=486 | - | N=1484 | N=936 |
| No adol. – no early adult. | 1 | - | 1 | 1 |
| Yes adol. – no early adult. | **0.36 (0.13-0.99)** | - | 0.91 (0.68-1.21) | 0.86 (0.19-3.81) |
| No adol. – yes early adult. | 0.78 (0.39-1.58) | - | 1.74 (0.61-4.95) | 1.08 (0.70-1.66) |
| Yes adol. – yes early adult. | 0.66 (0.29-1.51) | - | 0.73 (0.46-1.14) | 1.14 (0.54-2.39) |
| RERI<sup>a</sup> (95% CI) | 0.52 (-0.26-1.30) | - | -0.92 (-2.77-0.94) | 0.20 (-1.31-1.72) |
| **SMOKING** |  |  |  |  |
| **Women** | N=652 | N=268<sup>c</sup> | N=1922 | - |
| No adol. – no early adult. | 1 | 1 | 1 | - |
| Yes adol. – no early adult. | 1.49 (0.65-3.42) | **3.07 (1.29-7.35)** | 1.32 (0.97-1.79) | - |
| No adol. – yes early adult. | **0.33 (0.16-0.66)** | 1.86 (0.59-5.85) | 1.24 (0.81-1.91) | - |
| Yes adol. – yes early adult. | 1.08 (0.65-1.80) | 1.93 (0.97-3.85) | 0.93 (0.66-1.30) | - |
| RERI<sup>a</sup> (95% CI) | 0.26 (-1.06-1.58) | -2.01 (-5.39-1.38) | -0.63 (-1.35-0.09) | - |
| **Men** | N=488 | - | N=1421 | N=950 |
| No adol. – no early adult. | 1 | - | 1 | 1 |
| Yes adol. – no early adult. | 0.65 (0.20-2.11) | - | 1.03 (0.71-1.50) | 0.58 (0.08-4.53) |
| No adol. – yes early adult. | 0.96 (0.49-1.85) | - | 0.97 (0.52-1.83) | 0.90 (0.57-1.41) |
| Yes adol. – yes early adult. | 0.75 (0.35-1.61) | - | 1.26 (0.86-1.84) | 2.02 (0.79-5.12) |
| RERI<sup>a</sup> (95% CI) | 0.14 (-0.92-1.21) | - | 0.25 (-0.56-1.07) | 1.53 (-0.67-3.73) |

Abbreviations: TAM: Stress, Development and Mental health, TAM cohort; IDA: Individual Development and Adaptation; FT16: FinnTwin16;
NFBC1966: Northern Finland Birth Cohort 1966, GHQ-12: General Health Questionnaire; HED: heavy episodic drinking
a) Adjusted: Parental socioeconomic position (SEP) and psychological symptoms in adolescence/early adulthood
b) Adjusted only for parental SEP
c) Psychological symptoms measured in adolescence used as adjustment (not available in early adulthood)

### Combined associations of heavy episodic drinking and smoking

When combined associations between HED and smoking were examined by age phase, heterogeneity was shown in the association between engaging in both substances and distress in women in both age phases (adolescence I^2^ 67%, p=0.03, early adulthood *I*^*2*^ 68%, p<0.001). Again, the Swedish cohort showed higher ORs than the Finnish ones (combined substance use associated with distress in the IDA cohort in adolescence (OR 2.18, 95% CI 1.02-4.65) and in early adulthood (OR 2.60, 95% CI 1.14-5.95)). The associations in men were consistently non-significant, with some indication of heterogeneity in adolescence in engaging in both substances (*I*^*2*^ 68%, p=0.046). In the additive interaction analyses between HED and smoking on midlife distress, no interactions were found in either age phase (Table 4).

**Table 4.** Additive interaction analyses (and generalized mixed models in FT16) of heavy episodic drinking and smoking in adolescence and early adulthood and midlife psychological distress.

|  | TAM GHQ-12 | IDA GHQ-12 | FT16 GHQ-12 | NFBC1966 GHQ-12<sup>b</sup> |
| --- | --- | --- | --- | --- |
| ADOLESCENCE |  |  |  |  |
|  | N=727 | N=270 | N=2099 | N=3679 |
| **Women** | OR (95%CI) | OR (95%CI) | OR (95%CI) | OR (95%CI) |
| No HED – no smoking | 1 | 1 | 1 | 1 |
| Yes HED – no smoking | 0.71 (0.40-1.27) | 0.53 (0.06-4.45) | 1.09 (0.80-1.48) | 1.28 (0.96-1.72) |
| No HED – Yes smoking | 1.10 (0.57-2.14) | 1.76 (0.88-3.52) | 1.01 (0.69-1.48) | 1.62 (0.90-2.89) |
| Yes HED – Yes smoking | 1.11 (0.66-1.87) | **2.18 (1.02-4.65)** | 0.93 (0.62-1.40) | 0.94 (0.60-1.46) |
| RERI<sup>a</sup> (95% CI) | 0.30 (-0.66-1.26) | 0.89 (-1.01-2.79) | -0.17 (-0.79-0.44) | -0.96 (-2.05-0.12) |
| **Men** | N=588 | - | N=1613 | N=3086 |
| No HED – no smoking | 1 | - | 1 | 1 |
| Yes HED – no smoking | 0.82 (0.39-1.71) | - | 1.34 (0.93-1.95) | 1.24 (0.83-1.85) |
| No HED – Yes smoking | 1.17 (0.53-2.60) | - | 0.98 (0.60-1.58) | 1.01 (0.45-2.22) |
| Yes HED – Yes smoking | 0.61 (0.29-1.28) | - | 0.78 (0.47-1.29) | 1.61 (0.91-2.84) |
| RERI<sup>a</sup> (95% CI) | -0.38 (-1.54-0.79) | - | -0.54 (-1.31-0.23) | 0.37 (-0.92-1.65) |
| EARLY ADULTHOOD |  |  |  |  |
| **Women** | N=649 | N=216<sup>c</sup> | N=1956 |  |
| No HED – no smoking | 1 | 1 | 1 | - |
| Yes HED – no smoking | 1.75 (0.84-3.66) | 1.67 (0.62-4.50) | 1.06 (0.71-1.59) | - |
| No HED – Yes smoking | 0.74 (0.46-1.19) | 1.87 (0.72-4.82) | 0.97 (0.77-1.24) | - |
| Yes HED – Yes smoking | 0.48 (0.22-1.07) | **2.60 (1.14-5.95)** | 1.00 (0.75-1.34) | - |
| RERI<sup>a</sup> (95% CI) | -1.00 (-2.39-0.38) | 0.07 (-2.38-2.53) | -0.04 (-0.57-0.50) | - |
| **Men** | N=489 | - | N=1484 | N=947 |
| No HED – no smoking | 1 | - | 1 | 1 |
| Yes HED – no smoking | 0.63 (0.28-1.41) | - | 1.24 (0.67-2.28) | 1.32 (0.80-2.16) |
| No HED – Yes smoking | 0.94 (0.48-1.89) | - | 0.85 (0.62-1.18) | 1.49 (0.73-3.04) |
| Yes HED – Yes smoking | 0.76 (0.36-1.64) | - | 1.05 (0.73-1.51) | 1.11 (0.63-1.93) |
| RERI<sup>a</sup> (95% CI) | 0.20 (-0.71-1.11) | - | -0.04 (-0.84-0.76) | -0.69 (-1.97-0.58) |

Abbreviations: TAM: Stress, Development and Mental health, TAM cohort; IDA: Individual Development and Adaptation; FT16: Finn Twin16; NFBC1966: Northern Finland Birth Cohort 1966, GHQ-12: General Health Questionnaire; HED: heavy episodic drinking
a) Adjusted: Parental socioeconomic position (SEP) and psychological symptoms in adolescence/early adulthood
b) Adjusted only for parental SEP
c) Psychological symptoms measured in adolescence used as adjustment (not available in early adulthood)

## Discussion

This multicohort study looked for consistency in the associations between adolescent/early adulthood heavy episodic drinking and daily cigarette smoking with midlife psychological distress. A meta-analysis, consistent with findings from the four individual cohorts, found no clear evidence that heavy episodic drinking or smoking in adolescence and early adulthood were associated with psychological distress in midlife. This is somewhat inconsistent with our hypothesis and previous studies that have found longitudinal individual associations of HED/smoking and later psychological distress (Fergusson et al., 2009; McCambridge et al., 2011). Two issues may explain this discrepancy. First, mainly the previous evidence is based on diagnostic measures, emphasizing the more severe end of the symptom and substance use spectrum. From the public health perspective, a wider scope is relevant. Previous studies examining sub-clinical measures have usually followed up only until early adulthood. It may be that stronger associations found in previous studies are due to shorter follow-up times (proximity of predictors and outcomes).

Second, publication bias may distort the conclusions if more significant findings are selectively published. In the current study, data selection and study design were preplanned, reducing the tendency to find more positive findings. This procedure highlights the benefits of gaining more robust conclusions. We found rather similar findings across cohorts, increasing the confidence in these findings, and our key conclusions hold, despite some heterogeneity in the individual studies.

Regarding the role of different age phases, we hypothesized that the associations are stronger in adolescence compared to early adulthood, and the strongest when substance use continues from adolescence into early adulthood. Previous findings on the role of age in these associations have varied. Lee at al. (2022) highlighted that continued smoking is most detrimental for later mental health, and quitting smoking reduces the risk (see also Mathers et al., 2006). On the contrary, Trim et al. (2007) found that adolescent substance use is a risk for later mental health over and above the association with persistent use. All in all, smoking has previously mainly been shown to be a risk factor, as was the case also in this study in Swedish women. However, in early adulthood, opposite findings were found in women of the TAM cohort. In the adjusted analyses, early adulthood daily smoking was associated with less psychological distress in midlife. However, no such results were found among other cohorts or among men, which might suggest that gender and cohort-specific factors play a role in these associations.

These results underscore the complexity of the association between early substance use and subsequent mental health outcomes. It’s possible that the association of early substance use on later mental health diminished over time, or that the influence of other life experiences might mitigate this association (Lee et al., 2022, p. 202). In this multicohort design, we were not able to examine in detail the longitudinal trajectories of substance use extending beyond early adulthood. The problem is that co-occurring mental health and substance use problems are rather rare in adolescence (Berg and Kiviruusu, 2024), and identification of those at risk for later mental health problems is not easy, because many substance users in these age groups do not drift to harmful paths. Especially, in the Nordic context, HED in adolescence and early adulthood has been very typical in these age cohorts (Härkönen and Mäkelä, 2011; Kraus et al., 2015), while alcohol use and cigarette smoking are decreasing in younger cohorts (Ruokolainen et al., 2024). These cohorts were born in two countries between the 1950s and 1970s. We found no clear pattern regarding older cohorts having stronger associations than the younger ones. Investigating in more detail whether findings are similar across different contexts (Finland vs. Sweden, older vs. younger cohort) would require more cohorts to be included.

We expected a greater risk for psychological distress when both behaviors co-occur, which was found in Swedish women, but the findings suggested that the combined effect of HED and smoking in adolescence and early adulthood on midlife psychological distress did not exceed their individual effects. Particularly, women’s smoking was more common in the Swedish cohort compared to Finnish cohorts, but meta-analyses with cohorts from several countries should be conducted to examine country differences more specifically.

### Methodological considerations

In addition to the possibility to use several independent datasets, the main advantages of this study are prospective data collection, long follow-up times, and good participation rates. Despite several benefits of a multicohort study (e.g. generating a single estimate with greater precision, possibility to explore heterogeneity across different settings), in this approach, original data is not used in a way that it was necessarily designed for (O’Connor et al., 2022). For example, due to the harmonisation process, the measures used were dichotomized, and some nuances of the phenomenon may have been lost (e.g. occasional smoking). Another limitation is measurement bias; while the outcome (GHQ-12) was nearly the same in all cohorts (small differences in the scales), there were differences in HED measures (drunkenness vs. grams of alcohol). However, in the Finnish context, the concept of self-perceived drunkenness has been shown to correspond well with more objective measures (Lintonen and Rimpelä, 2001). Additionally, smokeless tobacco use is an increasingly relevant form of use (Ruokolainen et al., 2024), but we could not take this type of use or other drugs into account. The potential control variables varied between cohorts, and only a limited set was available across cohorts. NFBC1966 differs from other cohorts because adolescent substance use was measured at age 14, compared to age 16 in other cohorts. As a sensitivity analysis (data not shown), we examined adolescent substance use at age 15 in the TAM, but the associations remained statistically non-significant.

Regarding interpretation of meta-analysis, it should be taken into account that there were differences in sample sizes and attrition between cohorts. Some associations showed high *I*^2^ with non-significant Q., reflecting a small number of cohorts. The *I*^2^ might be high due to random variation rather than true heterogeneity.

## Conclusions

Using extensive follow-up and robust methodology, our findings did not provide clear evidence for individual or combined associations between heavy episodic drinking and cigarette smoking during adolescence and early adulthood with midlife psychological distress. These results were overall consistent across all four cohorts examined. The lack of associations challenges the prevailing notion that early-life substance use is a strong predictor of later-life psychological distress, particularly when considering subclinical measures of distress symptoms and risky health behaviors. This suggests that the relationship between substance use and mental health is complex and highlights the need for further research using diverse measures and longitudinal designs to fully understand these dynamics.

## Acknowledgements

We wish to thank all cohort members, researchers, and personnel in the cohort studies who participated in the data collections.

## Data availability statement

The information on the datasets analyzed during the current study is available on the following web pages www.thl.fi/en/tam (TAM cohort), http://www.helsinki.fi/fi/tutkimusryhmat/kaksostutkimus (Finntwin16), www.oulu.fi/nfbc (NFBC1966), and http://www.oru.se/english/research/projects/bsr/the-ida-program/ (IDA). The data underlying this article cannot be shared publicly due to legal restrictions and the nature of the data (individual-level data). The data are available upon request. Data requests are reviewed at the Finnish Institute for Health and Welfare (THL) (for TAM) and the Institute for Molecular Medicine Finland (FIMM) Data Access Committee (DAC) (for FinnTwin16) for authorized researchers who have IRB/ethics approval and an institutionally approved study plan. For more details, please contact THL and the FIMM DAC (fimm-dac@helsinki.f). NFBC1966 data are available from the University of Oulu, Infrastructure for Population Studies. Permission to use the data can be applied for research purposes via an electronic material request portal. In the use of data, we follow the EU general data protection regulation (679/2016) and the Finnish Data Protection Act. The use of personal data is based on a cohort participant’s written informed consent in their latest follow-up study, which may cause limitations to its use. Please, contact the NFBC project center (NFBCprojectcenter(at)oulu.fi) and visit the cohort website (www.oulu.fi/nfbc) for more information.

## Ethics statement

Ethical approval was given for each cohort from relevant ethical committees/boards. TAM cohort was approved by the Institutional review board (IRB) of The Finnish Institute for Health and Welfare (THL). FT16 was approved by the Ethical Committee of the Department of Public Health, University of Helsinki, Helsinki University Central Hospital ethical committee, and by the Institutional Review Board of Indiana University. NFBC1966 was approved by the Northern Ostrobothnia Hospital District Ethical Committee 94/2011 (12.12.2011). The IDA project has been examined by ethical committees on several different occasions. All participants gave informed consent to participate.

