## Supplementary information for "The individual and joint associations of alcohol use and cigarette smoking in adolescence and early adulthood with psychological distress in midlife – a multicohort study"

Supporting information

Description of study cohorts

The data comprise four prospective longitudinal studies, three from Finland and one from Sweden (Supplementary Table 1). The Finnish studies are the ‘TAM cohort’ (TAM), ‘FinnTwin16’ (FT16) and ‘Northern Finland Birth Cohort 1966’ (NFBC1966). TAM consists of pupils (N=2194) who attended the last year of compulsory school in 1983 in a city in Southern Finland. The participants completed questionnaires at ages 16, 22, 32, 42, and 52 (in 1983, 1989, 1999, 2009, and 2019). FT16 is a population-based study of five consecutive birth cohorts (1975-1979) of Finnish twins (N= 5563). Participants completed questionnaires at ages 16, 17, 18, 24, and 35 (in 1991-1995, 1992-1996, 1993-1997, 2000-2002, and 2010-2012). NFBC1966 was set with an expected date of birth in 1966 in the Oulu and Lapland area, comprising of 12,055 mothers and 12,231 children. The children have been followed from various sources, including questionnaires at ages 14, 31, and 46 (subsample of men also at age 24) (in 1980, 1997, and 2012).

The Swedish study is the ‘Individual Development and Adaptation’ (IDA), which consists of pupils (N=1031) who attended 3^rd^ grade in Örebro, Sweden in 1965. The participants have been followed up at ages 10, 13, 15, 16, 17, 18, 19, 26, 43 (only women), 47, and 49 (only women) (in 1965, 1968, 1970, 1971, 1972, 1973, 1974, 1981, 1998, 2002, 2004).

Supplementary Table 1. Information on TAM cohort, FinnTwin16 cohort, Northern Finland Birth Cohort 1966 and IDA project

| **Cohort** | **Area** | **Birth year** | **Adolescence** | | **Youth** | | **Early adulthood** | | **Midlife** | |
| --- | --- | --- | --- | --- | --- | --- | --- | --- | --- | --- |
|  |  |  | **Year** | **Age** | **Year** | **Age** | **Year** | **Age** | **Year** | **Age** |
| TAM  N=2194 | Tampere | 1967 | 1983 | 16 | 1989 | 22 | 1999 | 32 | 2009  2019 | 42  52 |
| FT16  N=5563 | Finland | 1975-1979 | 1991-1995  1992-1996 | 16  17 | 1993-1997  2000-2002 | 18  24 | 2010-2012 | 35 |  |  |
| NFBC1966  N=12058 | Oulu and Lapland | 1966 | 1980 | 14 | 1990 [men only (N=2500)] | 24 | 1997 | 30-31 | 2012 | 45-46 |
| IDA  N=1031 | Örebro | 1955 | 1970  1971 | 14  15 | 1981 | 26 |  |  | 1998 (women)  2002  2004 (subsample of men) | 43  47  49 |

Abbreviations: TAM: Stress, Development and Mental health, TAM cohort; IDA: Individual Development and Adaptation; FT16: FinnTwin16; NFBC1966: Northern Finland Birth Cohort 1966

In general, the participation rates have been high. In TAM, 53.0% of the original participants participated at age 52. In FT16, of those who were contacted, 72.0% participated at age 35. In the NFBC1966, questionnaire data at age 46 were received from 67% of those alive and address known in Finland. In IDA, the participation rate was 82% for women at age 49 and the same for men at age 47-48.

Supplementary Table 2. Original and harmonised variables in the TAM cohort, FinnTwin16 (FT16) cohort, Northern Finland Birth Cohort 1966 (NFBC1966) and IDA project

|  | **TAM** | **IDA** | **FT16** | **NFBC1966** | **Harmonised**  **variable** |
| --- | --- | --- | --- | --- | --- |
| **Outcome** |  |  |  |  |  |
| Midlife mental health | GHQ-12 | GHQ-12 | GHQ-12 | GHQ-12 | GHQ-12 |
| Age | 42 | 49 | 35 | 46 |  |
| Survey question | Next, we are asking how you have perceived yourself and your life over the last few weeks. | Have You in the past weeks... | Next, we would like to ask about your health and performance over the past month. Please answer all the questions by selecting the option that best describes your situation. Compare your well-being over the past month to what it usually is. | Have you lately… |  |
|  | 1. Have you recently been able to concentrate  on what you’re doing?  2. Have you recently felt capable of making  decisions about things?  3. Have you recently been able to face up  your problems?  4. Have you recently lost much sleep over  worry?  5. Have you recently felt constantly under  strain?  6. Have you recently felt you couldn’t  overcome your difficulties?  7. Have you recently been feeling unhappy  and depressed?  8. Have you recently been losing confidence  in yourself?  9. Have you recently been thinking of  yourself as a worthless person?  10. Have you recently felt you were playing a  useful part in things?  11. Have you recently been able to enjoy your  normal day-to-day activities?  12. Have you recently been feeling reasonably  happy, all things considered? | 1. …been able to concentrate on what you’re doing?  2. … had trouble sleeping because of anxiety?  3. … felt that you have an important role in what happens?  4. … felt capable of making decisions?  5. … felt very pressured?  6. … been able to cope with the daily problems?  7. … been feeling pretty happy considering the circumstances?  8. … felt that you cannot get over difficulties?  9. … been feeling unhappy and low-spirited?  10. … Begun to lose your confidence?  11. … begun to think of yourself as a worthless person?  12. … been able to enjoy activities of daily life? | 1. Have you recently been able to concentrate on whatever you're doing?  2. Have you recently felt capable of making decisions about things?  3. Have you recently been able to face up to your problems?  4. Have you recently lost much sleep over worry?  5. Have you recently felt constantly under strain?  6. Have you recently felt that you couldn't overcome your difficulties?  7. Have you recently been feeling unhappy and depressed?  8. Have you recently been losing confidence in yourself?  9. Have you recently been thinking of yourself as a worthless person?  10. Have you recently felt that you are playing a useful part in things  11. Have you recently been able to enjoy your normal day-to-day activities?  12. Have you recently been feeling reasonably happy, all things considered? | 1. …been able to concentrate on what you’re doing?  2. … felt capable of making decisions?  3. … been able to cope with the daily problems?  4. recently lost much sleep over worry?  5. …felt constantly under strain?  6. felt that you couldn't overcome your difficulties?  7. Felt unhappy and depressed?  8. lost self-confidence?  9. … begun to think of yourself as a worthless person?  10. been able to enjoy your normal day-to-day activities?  11. been feeling pretty been feeling reasonably happy, all things considered?  12. felt that your involvement is… |  |
| Values | 1-3   - Better than usual - As good as usual - Worse than usual - Much worse   4-9   - Not at all - Not more than usual - Somewhat more than usual - Much more than usual   10-12   - More than usual - As much as usual - Less than usual - Much less than usual | - Never - Some-times - Quite often - Always | 1-4   - Better than usual - Same as usual - Less than usual   5-12   - Not at all - No more than usual - Rather more than usual - Much more than usual | 1-3   - Better than usual - As good as usual - Worse than usual - Much worse   4-9   - Not at all - Not more than usual - Somewhat more than usual - Much more than usual   10-11   - More than usual - As much as usual - Less than usual - Much less than usual   12   - More usefull than usually - As usefull as always - Less usefull than usually - Much less usefull than usually |  |
| **Predictors** |  |  |  |  |  |
| Adolescent heavy episodic drinking (HED) | HED | HED | HED | HED | HED |
| Age | 16 | 16 | 16 | 14 |  |
| Survey question | During the past spring term, have you consumed alcohol so that you have been drunk? | Have You drunk so much beer, spirits or wine that You felt drunk? | …and how often do you get really drunk? | Have you been drunk? |  |
| Values | - no - yes, how many times? | - Never - 1 time - 2-3 times - 4-10 times - more than 10 times | - once a week or more - about 1-2 times a month - less often than that - never | - never - once slightly - twice or more times slightly - once very much - 2-4 times very much - several times very much | - No experience of HED or only a few times/less than monthly HED - Several experiences of being drunk/HED monthly or more often |
| Early adulthood HED | HED | HED | HED | HED | HED |
| Age | 22 | 26 | 25 | 24 (only men) |  |
| Survey question | And how often do you consume alcohol until heavily drunk? | How often do You drink alcohol? Folköl, pilsner, Starköl, Wine, Liquor How much alcohol do You drink the most at any one time? Enter the amount of different types - beer, wine, liquors - which You drink at such a time. | At present, how often do you within one occasion use more than five bottles of beer, or more than  a bottle of wine, or more than half a bottle of hard liquor (or a corresponding amount of alcohol)?  How often do you use alcohol to get drunk? | How often do you use alcohol so that you are drunk to relax or celebrate? |  |
| Values | - once or more times a week - about 1–2 times a month - less often - never | HED=60 grams or more on one occasion=5 or more drinks   - Every day - About once a week - About once a month - About once a year - Never | - daily - about twice a week - about once a week - a couple of times a month - about once a month - about once in two months - 3-4 times a year   8 once a year or less frequently  9 never  10 I don’t use alcohol at all | - never - a few times a year (e.g. on special occasions or holidays) - once or twice a month - about once a week - more than once a week | - Never or less than monthly - Monthly or more often |
| Adolescent smoking | Daily smoking | Daily smoking | Daily smoking | Daily smoking | Daily smoking |
| Age | 16 | 15-19 | 16 | 14 |  |
| Survey question | Which of the following options best describes your current smoking habits? | Please mark for each 5-year period how many cigarettes You smoked on a daily basis, in average. | which of the following best describes your present smoking  habits? | Smoking… |  |
| Values | - I smoke once a day or more often - I smoke once a week or more often, but not every day - I smoke less often than once a week - I have taken a break - I do not smoke |  | - I smoke once or more daily - I smoke once or more a week, but not every day - I smoke less often than once a week - I am trying to or have quit smoking - I have never smoked | - I have never tried - I tried once - I have tried twice or more - I smoke occasionally - I smoke about twice a week - I smoke 1-5 cigarettes daily - I smoke 6-10 cigarettes daily - I smoke more than 10 cigarettes daily | - No smoking or smoking but not every day - Daily smoking |
| Early adulthood smoking | Daily smoking | Daily smoking | Daily smoking | Daily smoking | Daily smoking |
| Age | 22 | 25-29 | 25 | 24 (only men) |  |
| Survey question | Which of the following options best describes your current smoking habits? | Please mark for each 5-year period how many cigarettes You smoked on a daily basis, in average. | Which of the following describes your smoking habits best? | Do you smoke nowdays? |  |
| Values | - I smoke once a day or more often - I smoke once a week or more often, but not every day - I smoke less often than once a week - I have taken a break - I do not smoke |  | - I smoke at least 20 cigarettes a day - I smoke 10-19 cigarettes a day - I smoke at most 9 cigarettes a day - I smoke once a week or more often but not daily - I smoke less than once a week - I have quit smoking - I have never smoked | - yes regularly (at least one cigarette per day) - yes, occasionally (less than one cigarette per day) - I don’t smoke | - No smoking or smoking but not every day - Daily smoking |
| Control variables |  |  |  |  |  |
| Adolescent mental health symptoms | Adolescent mental health symptoms | Adolescent mental health symptoms | Adolescent mental health symptoms |  | Adolescent mental health symptoms |
| Age | 16 | 15 | 16 |  |  |
| Survey question | During this spring term, have you experienced any of the following symptoms, and how often?  Circle the most appropriate option on each row. | Do you feel upset and bluesy without knowing the reason?  How often do you feel angry and ill-tempered to your mates?  How often would you be restless and have difficulty in staying still?  How often do you have bad appetite?  Do you have nightmares?  How often in this school year do you  have difficulty in falling asleep?  How often has it happened in this  school year that you have slept  unsettled and been awake at nights?  Do you feel lethargic and uncomfortable? | During the past six months have you had any of the  following symptoms and if so, how often?  Circle the closest alternative for each symptom. | Self-reported measures not available. |  |
| Time | Spring term | About the time you went to grade 8 (Autumn + part of spring term) | Past six months |  |  |
| Symptoms |  | sad and depressed | I am happy most of the time. (reversed) |  | Sadness |
|  | nervousness/anxiety | restless and difficulties staying still | tension or nervousness |  | Anxiety |
|  | irritability | angry and ill-tempered to your mates | irritability or temper outbursts |  | Irritability |
|  | loss of appetite | Bad appetite |  |  | Appetite |
|  | fatigue  apathy or lack of energy | lethargic and heavy hearted | fatigue or weakness |  | Listlessness |
|  | nightmares | nightmares |  |  | Nightmares |
|  | sleeping difficulties (Trouble falling asleep or waking up during  the night) | Difficulties falling asleep  Restless sleep | sleeping disorders |  | Sleeping difficulties |
| Values | - Not at all - Occasionally - Quite often - Often or continuously | - never - about 1 night/term - ’’ 1 night/month - ’’ 1 night/week - several nights/week      - very often - quite often - sometimes - occasionally - seldom - several times/week - about 1 time/week - ’’ 1 time/month - ’’ 1 time/term - never - almost never - occasionally - sometimes - quite often - very often | - Seldom or not at all - About once a month - About once a week - Almost daily |  | Standardized continuous variable |
| Family SEP | Parental occupation + education | Parental education | Parental occupation + education |  | Family SEP |
| Age | 16 | 16 | 16 | 14 |  |
| Survey question | What is your father’s education? What is your mother’s education? (Participant’s report) | What education do You have? (Parents’ self-report) | Which schools have you taken? (Parent’s self-report) |  |  |
| Values | - primary education or primary education and vocational education - civic school or civic school and vocational education - matriculation exam, or matriculation exam and vocational education - university or higher education degree - I don't know | - Folkskola - Realskola - Folkhögskola - Training school - Handels- eller tekn. inst. - Gymnasium, seminarium - Academic education - Other education (eg in company) Indicate approximately how long the education took | - Primary education or - other education - Primary education and at least one year of high school or vocational school - Matriculation examination - Matriculation examination and one year or education - University degree - other |  |  |
| Parental occupation |  |  |  |  |  |
| Age | 16 (14) | 16 | 16 | 14 |  |
| Survey question | Participant’s report:  What is the profession or occupation of your father or stepfather? Indicate the name of the profession or occupation as precisely as possible, e.g., machine engineer, upper secondary school teacher, student.  What is the profession or occupation of your mother or stepmother? Indicate the name of the profession or occupation as precisely as possible, e.g., office worker, shopkeeper, housewife. | Parent’s report:  Profession (not available) | Parent’s report (mother’s + father’s questionnaire): What is your occupation, or if you are not working, your previous occupation? | Parent’s report:  Mother's current occupation (give your mother's occupation even if she is not working tight now)  Father's current occupation  Also parent’s occupation has been asked. |  |
| Values | - upper non-manual - lower non-manual - manual   (based mainly on father’s occupation, if not available on mother’s and further on education) |  | - upper non-manual - lower non-manual - manual   (based on father’s and mother’s and if not available on education) | 0 ?  1 ?  2 ?  3 ?  4 ?  5 ? | - high - intermediate - low |

Supplementary Table 3 Percentages (cut off 2/3) of GHQ-12 in women and men

|  | WOMEN | | | | MEN | | |
| --- | --- | --- | --- | --- | --- | --- | --- |
|  | TAM | IDA | FT16 | NFBC1966 | TAM | FT16 | NFBC1966 |
|  | % (n), GHQ>3 | % (n), GHQ>3 | % (n), GHQ>3 | % (n), GHQ>3 | % (n), GHQ≥3 | % (n), GHQ≥3 | % (n), GHQ≥3 |
| SMOKING  (adol. + early adulthood) | * |  |  |  |  |  |  |
| no+no | 28.6 (132) | 19.4 (36) | 28.6 (437) | - | 15.3 (46) | 20.2 (209) | 11.5 (72) |
| only adol. smoking | 38.5 (10) | 37.2 (16) | 35.4 (40) | - | 14.8 (4) | 20.6 (14) | 7.1 (1) |
| only early adult smoking | 12.0 (10) | 25.0 (5) | 36.5 (84) | - | 16.5 (15) | 21.3 (53) | 10.5 (30) |
| yes+yes | 34.1 (29) | 25.6 (43) | 28.3 (62) | - | 15.3 (11) | 25.5 (52) | 20.7 (6) |
| HED  (adol. + early adulthood) |  |  |  |  |  |  |  |
| no+no | 27.2 (130) | 17.5 (27) | 29.6 (350) | - | 17.1 (51) | 20.6 (104) | 11.0 (41) |
| only adol. HED | 28.2 (29) | 24.4 (10) | 29.0 (27) | - | 9.1 (5) | 26.1 (6) | 9.5 (2) |
| only early adult. HED | 27.5 (11) | 31.5 (29) | 29.5 (215) | - | 15.0 (12) | 20.8 (189) | 11.8 (55) |
| yes+yes | 34.2 (13) | 26.3 (10) | 31.8 (54) | - | 16.4 (9) | 20.9 (42) | 12.5 (10) |
| ADOLESCENCE |  |  |  |  |  |  |  |
| No HED – no smoking | 26.5 (139) | 19.8 (33) | 29.0 (514) | 14.9 (461) | 16.0 (64) | 20.6 (276) | 10.6 (285) |
| Only HED | 22.1 (17) | 6.7 (1) | 33.6 (50) | 18.2 (63) | 15.2 (10) | 23.4 (29) | 12.8 (30) |
| Only smoking | 30.6 (15) | 26.7 (27) | 32.5 (79) | 22.1 (15) | 19.6 (9) | 27.5 (53) | 10.6 (7) |
| Yes HED – Yes smoking | 32.9 (27) | 28.0 (21) | 29.3 (41) | 14.0 (24) | 15.1 (13) | 18.9 (24) | 16.0 (15) |
| EARLY ADULTHOOD |  |  |  |  |  |  |  |
| No HED – no smoking | 28.0 (127) | 20.3 (26) | 29.0 (326) | - | 16.0 (42) | 20.9 (93) | 9.9 (32) |
| Only HED | 45.5 (15) | 26.0 (13) | 29.6 (171) | - | 13.6 (9) | 20.2 (150) | 12.9 (43) |
| Only smoking | 24.2 (30) | 25.4 (18) | 34.2 (52) | - | 15.1 (14) | 22.6 (21) | 13.8 (12) |
| Yes HED – Yes smoking | 20.5 (9) | 30.1 (25) | 31.5 (105) | - | 17.1 (12) | 23.3 (95) | 10.6 (25) |

*p<0.05

** p<0.01

*** p<0.001
